# Decontamination of SARS-CoV-2 and other RNA viruses from N95 level meltblown polypropylene fabric using heat under different humidities

**DOI:** 10.1101/2020.08.10.20171728

**Authors:** Rafael K. Campos, Jing Jin, Grace H. Rafael, Mervin Zhao, Lei Liao, Graham Simmons, Steven Chu, Scott C Weaver, Wah Chiu, Yi Cui

**Affiliations:** Department of Microbiology and Immunology, University of Texas Medical Branch, Galveston, Texas, United States; Vitalant Research Institute, San Francisco, California, United States; 4C Air Inc., Sunnyvale, California, United States; Department of Physics, Stanford University, Stanford, California 94305, United States; Department of Molecular and Cellular Physiology, Stanford, California 94305, United States; Institute for Human Infections and Immunity, University of Texas Medical Branch, Galveston, Texas, United States; Department of Bioengineering, Stanford University, Stanford, California, United States; Division of CryoEM and Bioimaging, SSRL, SLAC National Accelerator Laboratory, Menlo Park, California, United States; Department of Materials Science and Engineering, Stanford University, Stanford, California, United States; Stanford Institute for Materials and Energy Sciences, SLAC National Accelerator Laboratory, Menlo Park, California, United States

**Keywords:** COVID-19, SARS-CoV-2, coronavirus, humidity, N95, decontamination, aerosol

## Abstract

In March of 2020, the World Health Organization declared a pandemic of coronavirus disease 2019 (COVID-19), caused by the severe acute respiratory syndrome coronavirus 2 (SARS-CoV-2). The pandemic led to a shortage of N95-grade filtering facepiece respirators (FFRs), especially for protection of healthcare professionals against airborne transmission of SARS-CoV-2. We and others have previously reported promising decontamination methods that may be applied to the recycling and reuse of FFRs. In this study we tested disinfection of three viruses including SARS-CoV-2, dried on a piece of meltblown fabric, the principal component responsible for filtering of fine particles in N95-level FFRs, under a range of temperatures (60-95°C) at ambient or 100% relative humidity (RH) in conjunction with filtration efficiency testing. We found that heat treatments of 75°C for 30 min or 85°C for 20 min at 100% RH resulted in efficient decontamination from the fabric of SARS-CoV-2, human coronavirus NL63 (HCoV-NL63), and another enveloped RNA virus, chikungunya virus vaccine strain 181 (CHIKV-181), without lowering the meltblown fabric’s filtration efficiency.

---

The coronavirus disease 2019 (COVID-19) has affected millions of people globally and caused far-reaching impacts on public health and on the global economy.^1^ The etiologic agent, severe acute respiratory syndrome coronavirus 2 (SARS-CoV-2), a member of the family *Coronaviridae*, genus *Betacoronavirus*, subgenus *Sarbecovirus*, emerged in 2019. Its transmission is thought to take place through droplets and subsequently formed aerosols, generated when infected people sneeze, cough, speak, sing and even breathe, or by contact between people or through fomites.^2-5^ Data from influenza virus research revealed that aerosols generated by people can be classified into coarse (>5 μm) or fine (<5 μm) particles, and while coarse particles settle within one hour, fine particles can remain suspended in the air for long-periods of time when in poorly ventilated indoor environments;^6-8^ in the case of SARS-CoV-2, it can remain infectious in fine particle aerosols for at least 16 hours.[CITE] The virus particles contained in these aerosols can be inhaled to infect cells of the upper or lower respiratory tracts.^6-7^ The use of particulate filtering facepiece respirators (FFRs) is a form of personal protection which can minimize the inhalation of small airborne particles^9^ by medical workers, or other professionals at high risk of infection by SARS-CoV-2. Recent systemic studies demonstrate that facial coverings, including FFRs and masks, may reduce COVID-19 spread by 85%^10^ and are a highly important mitigation measure.^5^ FFRs of N95 or other highly filtering grade (*e.g*. FFP2, KN95, DS/DL2 and KF94) are able to filter 75 nm particles (median diameter) with > 95% efficiency, which should provide sufficient protection against SARS-CoV-2, which measures ~120nm or larger in aerosol particles containing the virus.^11^

The SARS-CoV-2 pandemic has led to shortages of personal protective equipment (PPE), including N95-grade FFRs. To mitigate this issue, different methods were proposed to decontaminate these face masks and allow for safe re-utilization: heat,^12-15^ ultraviolet (UV) irradiation,^12, 16^ steam,^17^ ozone,^18^ vaporized hydrogen peroxide (VHP),^19^ chemical disinfectants,^12^ and autoclaving.^20^ Although some of these methods show promise, most can lead to a reduction in the meltblown’s filtration efficiency or alteration in the physical fit in the FFR,^12, 16^ making them potentially unsafe for repeated usage. In addition, the proposed decontamination methods that may not alter the physical properties of FFR (filtration efficiency and fit) have varying levels of efficacy. For example, mild UV irradiation, which is an accessible and promising method for FFR decontamination when used within certain dosages, may have poor penetration when many layers of material are present. This raises the concern that the virus may be protected from inactivation if it penetrates deep into the layers of the FFR.^21^ On the other hand, decontamination using moderate heat (< 100°C) has emerged as one of the most promising methods to decontaminate FFRs during the COVID-19 pandemic because it causes little damage to the FFR^12, 22^ and can be done in high throughput. While there is evidence that viruses can show considerable resistance to dry heat after being dried on surfaces,^23-24^ detailed data are not consistently available for all viral families. For example, one study found that when influenza virus was dried on a stainless-steel surface, heating was more effective at reducing viral infectivity when the humidity was high.^24^ Heating influenza virus at 60°C for 15 min at 25% relative humidity (RH) resulted in a decrese of 1 log_10_-fold in viral titers, whereas the same heating process at 50% RH reduced viral titers by a factor of 4 log_10_-fold.^24^ Although SARS-CoV-2 has been shown to be efficiently inactivated by heat in solution ^25^, a comparison of inactivation by dry or moist heat when the virus is dried on surfaces has not been carried out, although one study found that stability of SARS-CoV-2 is lower in high humidity at common environmental temperatures (≤38°C).^26^

To further address the relationship between humidity and virus susceptibility to heat inactivation, we tested heat inactivation protocols using three positive strand enveloped RNA viruses: SARS-CoV-2, human coronavirus NL63 (HCoV-NL63), and chikungunya virus, vaccine strain 181/25 (CHIKV-181). We tested infectivity of virus samples dried on meltblown fabric, before subjecting the samples to a treatment of temperatures (60-95°C) at either ambient (40-60% RH) or 100% RH.

## RESULTS

FFRs of N95 or equivalent grade are usually composed of many layers of polypropylene nonwoven fabrics. Among the most important layers for its protective functionality is one produced by the meltblown process (i.e. meltblown nonwoven fabric), in which high velocity air blows a molten polymer, forming filaments that extend in different orientations and entangle in a web shape. When used in FFRs, this layer is electrostatically charged to significantly increase fabric’s filtration capability for small particles, due to electrostatic adsorption ^12^ We first visualized the structure of the meltblown fabric using scanning electron microscopy (SEM). Images of the meltblown fabric tested exhibited microfibers with diameters in the range of 2-10 p,m which cross each other to form a three-dimensional porous structure (Figure 1A). The meltblown fibers are typically electrostatically charged to increase binding of particles, resulting in a much higher filtration efficiency without increasing the air resistance. To test the particle adsorption of the meltblown fibers, we used sodium chloride (NaCl) solution as an aerosol source (0.26 μm mass median diameter, 0.075 μm count median diameter) and loaded the NaCl aerosol onto the meltblown fabric. SEM images showed clear NaCl particles adsorbed onto the meltblown fibers (Figure 1 B and C).

**Figure 1.**
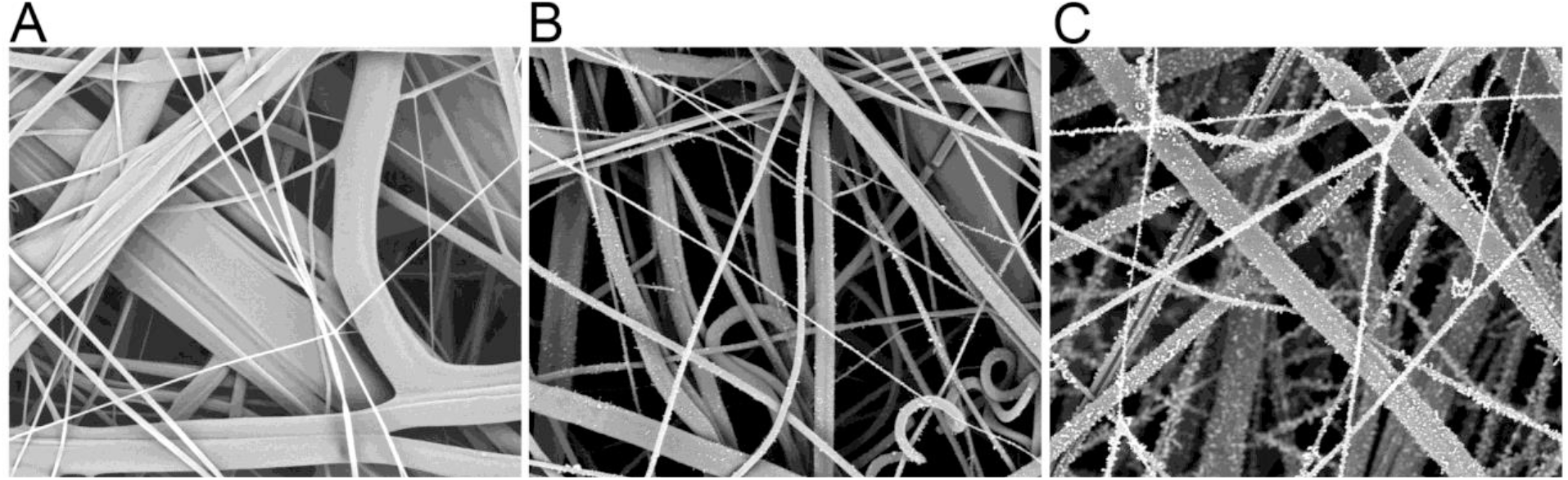
Scanning electron microscope (SEM) images of meltblown fabric before and after aerosol loading. Images of one piece of meltblown fabric (A) before and after loading with aerosol containing NaCl particles for 1 min (B) or 10 min (C). Particles trapped in the fabric can be observed in B and C.

As the primary filtering material for small particles in FFRs, meltblown fabric was used to give a worst-case scenario for how the filtration efficiency changes as respirators are treated under various temperatures and humidities. Thus, if the meltblown fabric is observed to be unaffected by the treatment, it is not expected that a whole FFR’s filtration properties would be affected. However, if the meltblown fabric’s filtration properties are changed, this does not necessarily indicate that a full FFR’s filtration properties would be affected, as the full respirator has other supporting materials and is made up by multilayers, which can make it more robust. In addition, we only considered how the meltblown’s filtration efficiency would change, but as FFRs also require proper fitting, it is possible that treatment conditions can alter the fit of FFRs. The data currently show that with respect to heat decontamination, it is unlikely to alter the fit of FFRs.^27^

We used a meltblown fabric of 20 g/m^2^ with initial efficiency around 95% (details given in the Methods section) to simulate how the filtration efficiency would change after application of heat for multiple treatment cycles. We first performed filtration efficiency testing after heat treatment of the meltblown fabrics under low humidity conditions (≤30%) to determine the upper limt of applicable heat. We observed no changein the filtration efficiency after treatment from 75 °C to 100 °C, with up to twenty treatment cycles hovering around 95% filtration efficiency. However, we observed that heating at 125 °C led to a sharp drop in the filtration efficiency at the fifth cycle, leading to a filtration efficiency of around 90%. In contrast to the filtration efficiency, we observed that the air resistance of the fabric (Figure 2B) did not vary when heat treatments of different temperatures were applied, which indicates that the physical structure and porosity of the material remained constant. Therefore, it is likely that high heat reduces the electrostatic charge on the meltblown fibers. This is not surpising, as polypropylene, the primary component of meltblown fabrics, has a melting point between 130 – 171 °C. Thus, as the temperature approaches the melting point, the crystalline structure can become more relaxed, and it is possible that this will affect the filtration properties of the fabric through charge relaxation or other means. We determined that the upper limit of low humidity heat may be <100 °C for treatment cycles ≤20 (cycle lengths are given in the Methods section for different temperatures). The data from these experiments is available in Supplementary Table S1.

**Figure 2.**
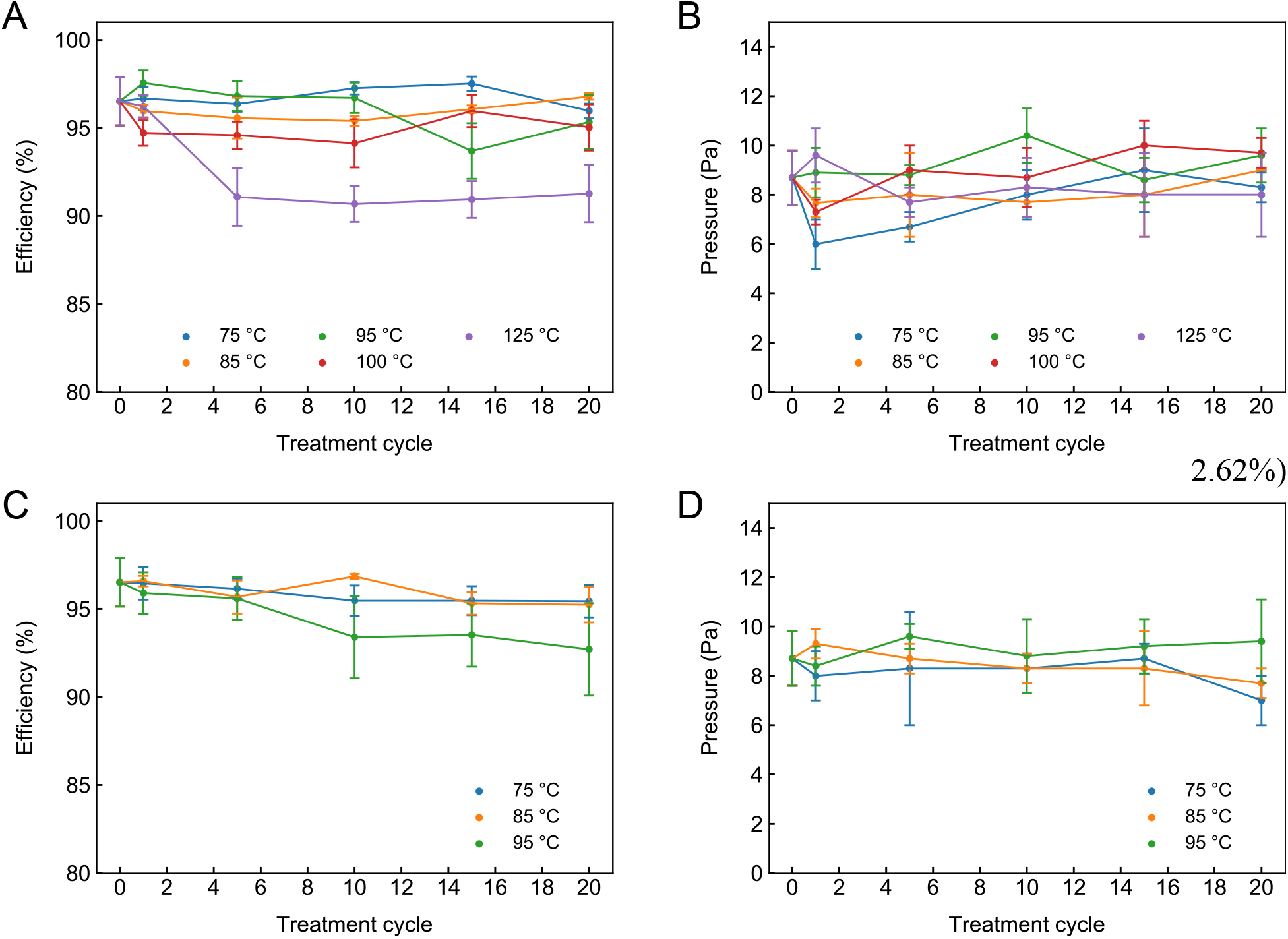
Filtration properties of meltblown fabric after multiple cycles of treatment at different temperatures. Meltblown fabric with filtration efficiency ≥95% was cut to approximately 15 cm × 15 cm pieces. Filtration efficiency and pressure drop were measured on an Automated Filter Tester 8130A. The flow rate for all measurements was 32 L/min, and NaCl was used as an aerosol source (0.26 μm mass median diameter, 0.075 μm count median diameter). Heat treatment under (A-B) ambient RH (40%) or (C-D) 100% RH were performed, and filtration efficiencies (A and C) and pressure drop (B and D) of the meltblown fabric were recorded.

As previously discussed, many viruses may have decreased viability in moist heat, so we also determined how the filtration efficiency would change under application of near 100% RH heat. As our previous work demonstrated that steam decreased the filtration efficiency,^12^ we chose to test the moist heat treatments at 75 °C, 85 °C, and 95 °C. Simulating 100% RH was performed by sealing the meltblown fabrics in polyethylene bags with 0.3 mL of water, not contacting the meltblown fabric (details in Methods section). For the samples tested at 75 °C and 85 °C, we observed no significant changes in filtration efficiency for twenty treatment cycles compared to the initial value (Figure 2C). For the samples tested at 95 °C, we observed that five cycles of temperature treatment did not affect the filtration efficiency (Figure 2C). However, by ten cycles, the samples tested at 95 °C decreased in filtering efficiency more than at the other two temperatures, though with a larger uncertainty as well (93.39% ± 2.33%) (Figure 2C). This filtration efficiency remains nearly constant and slightly decreases by twenty cycles (92.70% ± Figure 2C). As previous measurements showed that steam from boiling water also significantly decreased the filtration efficiency to values as low as ~80% by the tenth treatment cycle,^12^ the decay observed at 95 °C is not surprising and treatment cycles at this temperature should be limited to a minimum. It is possible that this decay is due to some adsorption of water, or other mechanisms which can decay the electrostatic charge, as the air resistance also did not change under these conditions (Figure 2D), similar to the case for the low humidity 125 °C samples.

Regarding the applicable temperatures, we can conclude that under low humidity (≤30% RH), temperatures ≤ 100 °C (twenty cycles) did not significantly degrade the filtration efficiency. Under high humidity (~100% RH) temperatures should be limited to ≤ 85 °C (twenty cycles) or ≤ 95 °C (five cycles) to preserse the filtration efficiencies found in the meltblown fabrics we tested (Figure 2 and Supplementary Table S1).

We next aimed to determine conditions of heat treatment and humidity that are efficient in inactivating SARS-CoV-2 loaded on the meltblown fabric. We mixed our virus stock 1:10 with either phosphate buffered saline (PBS) or bovine serum albumin (BSA) to a final concentration of 3g/L, to mimic the conditions of bodily fluids such as sputum,^28-30^ which contain higher protein concentrations than is found in the media used to create virus stocks; this is relevant because high protein concentrations are known to stabilize viruses, including SARS-CoV-2.^31^ We allowed droplets containing SARS-CoV-2 (backtiter 1 × 10^6^ PFU/sample) dry on top of a piece of meltblown fabric in a biosafety cabinet for 2 hours. These pieces were then added to a microcentrifuge tube that was incubated at ambient RH of 60% on a heat block at 25°C, 60°C, 75°C for 30 min, 85°C for 20 min, or 95°C for 5 min. The virus was then recovered by the addition of media followed by vortexing, from which we were able to recover approximately 1 × 10^5^ PFU in the 25°C control condition. In the absence of BSA, heating the fabric at 60°C for 30 min resulted in a 2 log_10_-fold reduction in viral titers in comparison to 25°C for 30 min, whereas treatment at 75°C for 30 min reduced viral titers by 3.5 log_10_-fold and heating at 85°C for 20 min or 95°C for 5 min reduced the virus by 5 log_10_-fold, bringing the titers below the limit of detection (LOD) of the assay (Figure 3A and Table 1). Our tests indicated that the addition of BSA stabilized the virus, as expected, and in these conditions the titers were reduced by 1 log_10_-fold at 60°C for 30 min, 3 log_10_ at 75°C for 30 min, 4 log_10_-fold at 85°C for 20 min, and 5 log_10_-fold at 95°C for 5 min. Strikingly, virus could be detected in all of the conditions tested, although the 95°C treatments reduced the titers greatly and the amount of virus detected neared the LOD (Figure 3A).

**Figure 3.**
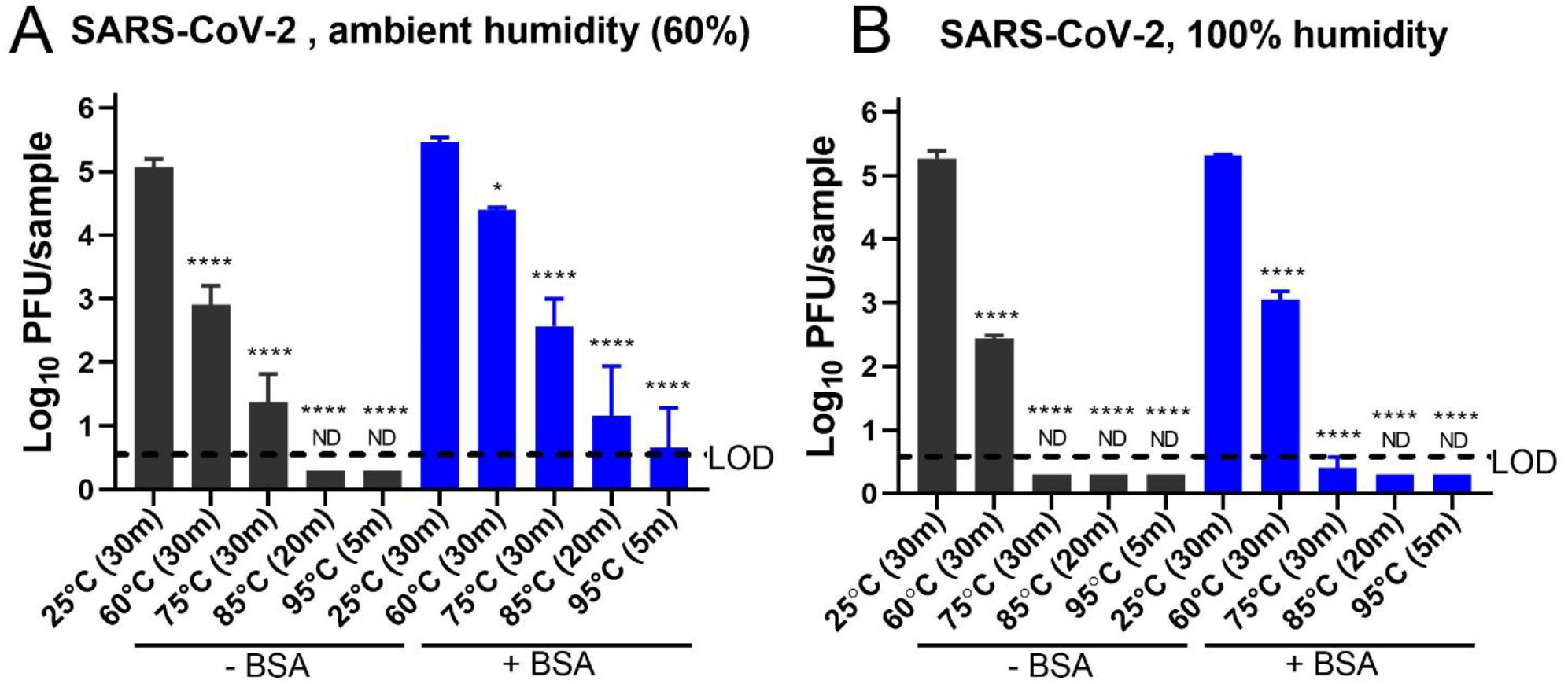
Heat inactivation of SARS-CoV-2 dried on a piece of meltblown fabric is more efficiently inactivated by 100% humidity. A. Heat treatment of SARS-CoV-2 dried on meltblown fabric at ambient humidity (approximately 60%). B. Heat treatment of SARS-CoV-2 dried on meltblown fabric at 100% humidity. Error bars represent SD. ND (not detected) are conditions in which each of the triplicates were below the LOD. Statistical significance was assessed by oneway ANOVA using Sidak’ multiple correction test. * p < 0.05, **** p < 0.0001.

**Table 1.** Reduction of SARS-CoV-2 titers by different heat inactivation conditions.

| Virus | Condition | Temperature | Reduction from 25°C in log<sub>10</sub> |
| --- | --- | --- | --- |
| SARS-CoV-2 | Ambient humidity (60%) - BSA | 60°C (30 min) | 2.16 +/- 0.23 |
|  |  | 75°C (30 min) | 3.69 +/- 0.32 |
|  |  | 85°C (20 min) | > 4.77 |
|  |  | 95°C (5 min) | > 4.77 |
|  | Ambient humidity (60%) + BSA | 60°C (30 min) | 1.07 +/- 0.06 |
|  |  | 75°C (30 min) | 2.89 +/- 0.31 |
|  |  | 85°C (20 min) | 4.3 +/- 0.55 |
|  |  | 95°C (5 min) | 4.8 +/- 0.44 |
|  | 100% humidity - BSA | 60°C (30 min) | 2.82 +/- 0.09 |
|  |  | 75°C (30 min) | > 4.97 |
|  |  | 85°C (20 min) | > 4.97 |
|  |  | 95°C (5 min) | > 4.97 |
|  | 100% humidity + BSA | 60°C (30 min) | 2.27 +/- 0.09 |
|  |  | 75°C (30 min) | 4.92 +/- 0.12 |
|  |  | 85°C (20 min) | > 5.02 |
|  |  | 95°C (5 min) | > 5.02 |

Air humidity is one factor that has generally been considered important for inactivation of dried viruses,^23-24^ and should therefore be taken into account when performing heat inactivation studies. We therefore tested whether SARS-CoV-2 dried on meltblown fabric could be inactivated by heat at 100% RH. To achieve 100% RH, we added 100 |il of water to the bottom of each tube with the meltblown fabric, without touching the fabric. Using these moist heat conditions, we observed improved inactivation; in the absence of BSA, we observed a 3 log_10_-fold decrease in viral titers when samples were incubated at 60°C for 30 min, 1 log_10_-fold more than in the ambient RH conditions (Figure 3B and Table 1). This also led to improved inactivation at higher temperatures, and no virus could be detected using temperatures from 75°C to 95°C (Figure 3B and Table 1). In the presence of BSA at 60°C for 30 min, there was a 2 log_10_-fold decrease in viral titers at 100% RH in comparison to a decrease of 1 log_10_-fold in the ambient humidity conditions (Figure 3B). When heated at 75°C for 30 min, we observed a decrease of almost 5 log_10_-fold in viral titers, with only one of three replicates being detectable (Figure 3B and Table 1).

To test whether our findings are generalizable to other coronaviruses, we tested the heat sensitivity of human coronavirus NL63 (HCoV-NL63) from the genus *Alphacoronavirus*, subgenus *Setracovirus*, and another (+)ssRNA enveloped virus. CHIKV-181, a member of the family *Togaviridae*, genus *Alphavirus*. These viruses were also dried on meltblown fabric, using the same protocol as for SARS-CoV-2. At 60°C or 75°C for 30 min in ambient RH (40% RH) HCoV-NL63 was reduced 0.5 or 1.5 log_10_-fold, respectively, whereas CHIKV-181 was reduced by a little over 1 log_10_-fold at both of these conditions (Figure 4A and B). At higher temperatures, the stabilities of HCoV-NL63 and CHIKV-181 were comparable, showing about 2.5 log_10_-fold reduction at 85°C for 20 min or 95°C at 5 min (Figure 4A and B). HCoV-NL63 and CHIKV-181 inactivation was done in ambient RH (40%) while SARS-CoV-2 inactivation was done in ambient RH (60%). This may explain the increased resistance of HCoV-NL63 and CHIKV-181 to heat inactivation compared with SARS-CoV-2. We next tested the heat stability of these viruses on meltblown fabric under 100% RH conditions, either without or with 3 g/L of BSA as a stabilizer. Under the conditions of 100% RH and without BSA, both HCoV-NL63 and CHIKV-181 were reduced by 2 log_10_ when heated at 60°C for 30 min (Figure 4C and D). When BSA was added, HCoV-NL63 was differentially more stabilized than CHIKV-181 (Figure 4C and D). After heating for 30 min at 60°C, CHIKV-181 titers were reduced by 2 log_10_-fold, whereas HCoV-NL63 titers were reduced by only 1 log_10_-fold; and after heat treatment for 30 min at 75°C, CHIKV-181 titers were reduced by 4 log_10_-fold in comparison to a reduction of 2 log_10_-fold for HCoV-NL63 (Figure 4C and D). CHIKV-181 was not detected following either the 85°C or 95°C treatments, whereas HCoV-NL63 titers were below the LOD at 85°C for 20 min but still detectable when treated with 95°C for 5 min (Figure 4C and D). Although a direct comparison was not done with SARS-CoV-2 the reductions observed for CHIKV-181 were comparable with SARS-CoV-2, and these viruses were slightly more sensitive to heat inactivation than HCoV-NL63 at 100% RH (Figures 4B and 4C and D). Taken together, these results suggest that moist heat is an effective method to decontaminate FFRs from SARS-CoV-2 and other RNA viruses.

**Figure 4.**
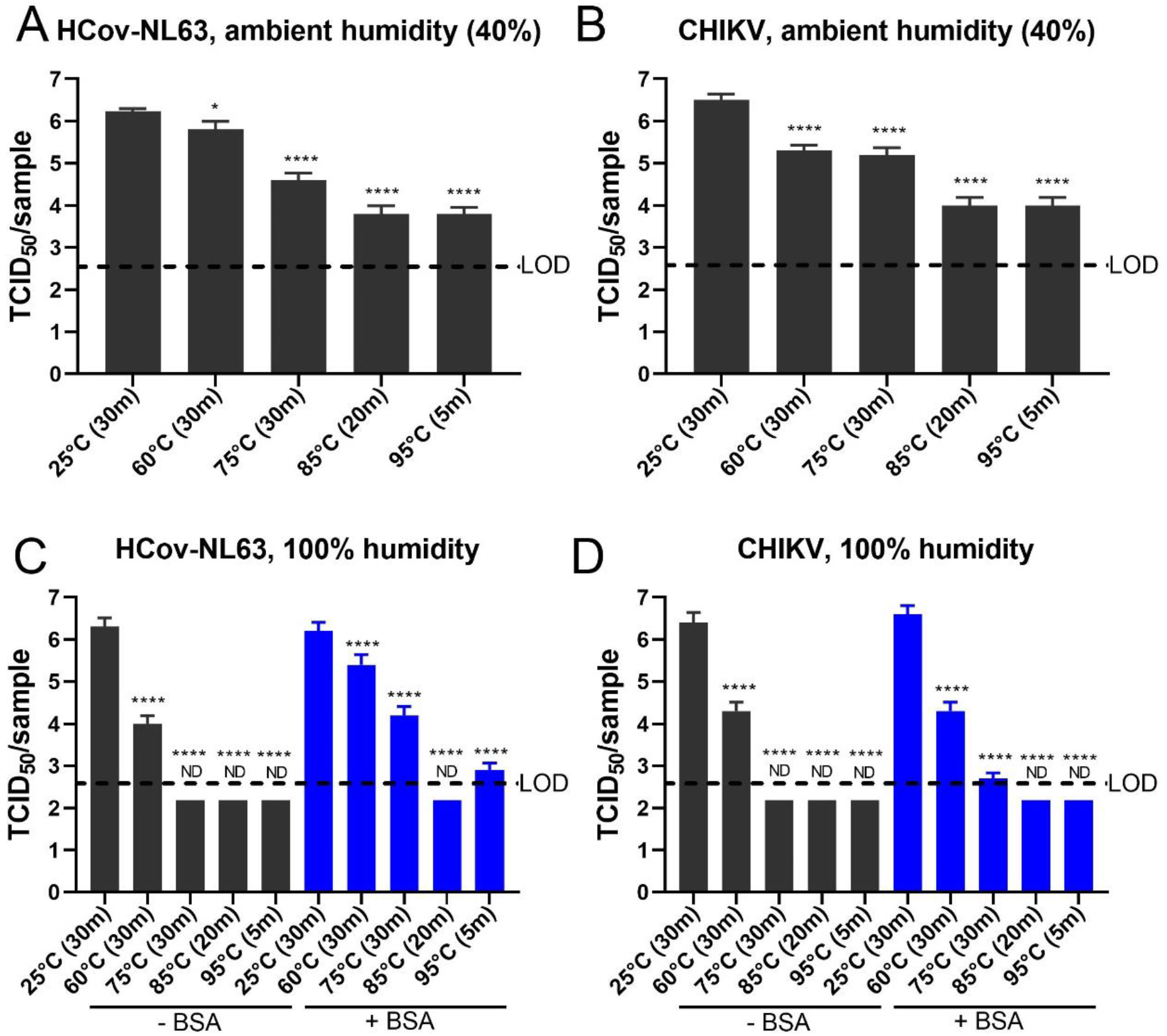
Heat inactivation of other (+)ssRNA viruses CHIKV-181 and HCoV_NL63 dried on a piece of meltblown fabric are more efficiently inactivated by 100% humidity. Heat inactivation of (A and C) HCoV-NL63 and (B&D) CHIKV-181181 dried on meltblown fabric at (A and B) ambient humidity (approximately 40%) and (C&D) 100% humidity. Error bars represent SD. ND (not detected) are conditions which all replicates were below the limit of detection (LOD). Statistical significance was assessed by one-way ANOVA using Sidak’ multiple correction test. **** p < 0.0001.

## CONCLUSIONS

COVID-19 is an exceptionally contagious disease that requires healthcare workers to take many precautions including the use of PPE to protect themselves. The shortage of PPE caused by the COVID-19 pandemic can be mitigated by decontamination of this equipment, allowing for safe reutilization. We tested different temperatures for heat inactivation of SARS-CoV-2 dried on a piece of meltblown fabric, a key component of N95 grade FFRs, under conditions of ambient or 100% RH. We found that temperatures of 75°C-85°C are able to efficiently inactivate the virus in 20-30 min under 100% RH, without lowering filtration efficiency. This humidity does not extensively impact filtration, as FFRs can undergo at least twenty cycles of 75°C for 30 min/cycle or 85°C for 20 min/cycle at 100% RH without losing filtration efficiency. Although there remains uncertainty regarding whether other pathogens can be effectively inactivated by moist humidity, this is a promising decontamination method in the context of the COVID-19 pandemic. Our work also revealed that humidity has a profound effect on heat inactivation of dried SARS-CoV-2, which should be taken into consideration for public policies to prevent infection by SARS-CoV-2.

## METHODS

### Air filtration efficiency measurement of meltblown fabric

Meltblown fabric with filtration efficiency ≥95% was procured from Guangdong Meltblown Technology Co., Ltd. It has a base weight of 20 g/m^2^. All samples were cut to approximately 15 cm × 15 cm. Filtration efficiency and pressure drop were measured on an Automated Filter Tester 8130A (TSI, Inc.) The flow rate for all measurements was 32 L/min, and a 2% NaCl solution was used to generate aerosol with 0.26 μm mass median diameter/0.075 μm count median diameter. Each uncertainty is determined from at least three measurements. Samples that were tested were not re-tested.

The conditioning and aging of samples under different temperatures and humidities was performed in an SH-642 environmental chamber. The chamber can control relative humidity to a minimum of 30% for temperatures <85 °C. Above 85 °C, the relative humidity is <30% but not measured. Relative humidities of 100% were simulated by placing the meltblown fabrics into sealed plastic bags with 0.3 mL of water. For 75 °C, the samples were aged for 30 min per cycle, for 85 °C the samples were aged for 20 min per cycle, for 95 °C the samples were aged for 15 min per cycle, for 100 °C the samples were aged for 10 min per cycle, for 125 °C the samples were aged for 10 min per cycle. All scanning electron microscope (SEM) images were taken on a Phenom Pro SEM, with 10 kV as the electron voltage.

### Cells and viruses

Vero E6 (ATCC® CRL-1586™) and MA-104 (<u>ATCC® CRL-2378.1</u>) cells were maintained in Dulbecco’s minimal essential media (DMEM, Gibco) and ATCC-formulated Eagle’s Minimum Essential Medium (EMEM, ATCC) supplemented with 5% fetal bovine serum (FBS, Atlanta Biologicals) and 1% penicillin/ampicillin (Gibco). Cell cultures were maintained in an incubator set to 37°C with 5% CO_2_. SARS-CoV-2 strain USA_WA1/2020 was obtained from the World Reference Center for Emerging Viruses and Arboviruses (WRCEVA) at passage 3 (in Vero cells) and amplified in Vero E6 cells to generate a working stock. A confluent flask was infected with SARS-CoV-2 at an MOI of 0.001 in 5ml of DMEM supplemented with 2% of FBS, 200 mg/ml of streptomycin, and 200 U/ml of penicillin.. Adsorption was allowed to proceed for 1 h in an incubator set to 37°C with 5% CO_2_, rocking every 15 min. The medium was then removed and 20 ml of DMEM supplemented with 2% of FBS and antibiotics. After 3 days of infection, when widespread cytopathic effect (CPE) was observed, supernatant was collected, spun for 5 min at 3,000 × g on a tabletop centrifuge, aliquoted and stored at -80°C. HCoV_NL63 strain and CHIKV-181 vaccine strain 181/25 were obtained from the Biodefense and Emerging Infections Research Resources Repository (BEI) and amplified in Vero E6 cells.

### Heat inactivation of viruses on meltblown fabric

Heat tests of SARS-CoV-2 inactivation on metlblown fabric were performed in a biosafety level 3 (BSL3) lab, on either an absolute humidity of 100% or an ambient with absolute humidity (60% +/− 1%, as measured during the experiment). Pieces of meltblown fabric were cut into 2 × 2 cm squares and 20 droplets of 5 μl each of virus stock (8 × 10^6^ PFU/ml) were dripped on top of the fabric. The virus was allowed to dry for 2 h at room temperature in the biosafety cabinet. The pieces of fabric with dried virus were then transferred to a microcentrifuge tube using sterilized forceps. For the ambient humidity experiments, the microcentrifuge tubes were empty, whereas for the 100% humidity experiments, 100 ^l of autoclaved deionized water was added to the bottom of the tube (the fabric did not touch the water). Tubes were then heated at the specified temperatures using heat blocks. After heat treatment, DMEM supplemented with 2% of FBS and 1% penicillin/ampicillin was added to each tube, 900 ^l in the 100% humidity conditions and 1 ml in the ambient humidity conditions. The tubes were vortexed for 30 s each and the virus was immediately titrated. Heat inactivation of HCoV_NL63 and CHIKV-181 181/25 on meltblown fabric was performed in a BSL2 lab at Vitalant Research Institute as described for SARS-CoV-2 above.

### Plaque assay

Vero E6 6-well plates were prepared the previous day by adding 800,000 cells per well. Samples to be titrated were serially diluted in DMEM supplemented with 2% of FBS and 1% penicillin/ampicillin. From each dilution, 250 |il were added to the Vero E6 monolayer, and samples were incubated with the virus for 1h in an incubator set to 37°C with 5% CO_2_, rocking every 15 min. After 1h, cells were overlayed with 4 ml of a solution containing 0.8% agarose, 4% FBS and antibiotics in minimum essential medium (MEM) without phenol red (Thermo Fisher). The plate was incubated for 2 days at 37°C with 5% CO_2_ and then 2 ml of 0.05% neutral red (Thermo fisher) were added on top of the overlay for 6 h. Neutral red was then removed and plaques were visualized in a biosafety cabinet using a light box and plaques were counted.

### TCID50 assay

Titers of HCoV_NL63 and CHIKV-181 181/25 were measured by TCID_50_ using MA-104 and Vero E6 cells respectively. Viruses were serially diluted in respective medium supplemented with 2% FBS prior to addition to cell monolayer in 96-well-plate. For each dilution, viruses were added to 10 replicate wells at 100 μl per well. The plates were incubated for 2 days (for CHIKV-181 181/25) or 6 days (for HCoV_NL63) at 37°C with 5% CO_2_ until clear cytopathic effect developed. Cells were stained with crystal violet and 50% endpoints were calculated with Reed and Muench method ^32^.

### Statistical analyses of plaque assays and TCID_50_ assays

To assess statistical differences in viral titers, one-way ANOVA with Sidak’s multiple comparison test was used to compare each condition to the control condition (25°C for 30 min). The LOD is the theoretical lowest amount of virus that can be detected in one replicate. For table 1 reductions are shown as the difference between the averages of the 25°C and each temperature of the same treatment condition. Error represents pooled standard deviation of the samples treated at 25°C and the samples treated at each temperature. Samples below the LOD were indicated as being more reduced than the difference between the 25°C condition and the LOD.

## Data Availability

All the data would be available upon request.

## ASSOCIATED CONTENTS

### SUPPORTING INFORMATION

The supporting information is available online. This includes the raw data measurements used for the filtration fficiency at various temperatures and humidities.

## AUTHOR CONTRIBUTIONS

Y.C., S.C and W.C. conceived the investigations. R.K.C., J.J., W.C., S.C., S.C.W. and Y.C. designed the experiments, R.K.C., J.J. and G.H.R. collected the virus disinfection data. M.Z. and L.L. collected the filtration efficiency results. R.K.C. and J.J.analyzed the data and interpreted results through discussions and contributions of W.C., S.C., S.C.W. and Y.C. R.K.C. and J.J. wrote the first version of the manuscript. The final version of the manuscript was written through contributions of R.K.C., J.J., W.C., S.C., S.C.W. and Y.C. All authors have given approval to the final version of the manuscript. R.K.C. and J.J. contributed equally.

## NOTES

The authors declare the following competing financial interest(s): Professors Steven Chu and Yi Cui are founders and shareholders of the company 4C Air, Inc. They are inventors on patent PCT /US2015/065608.

## ACKNOWLEDGMENT

We thank our colleagues, especially Nehad Saada and Dr. Sasha Azar, for their helpful suggestions.

